# Representing Injuries in Trauma Patients: Development and Evaluation of Embeddings for Injuries

**DOI:** 10.64898/2026.01.03.26343379

**Authors:** Kelvin Szolnoky, Jonatan Attergrim, Awais Ashfaq, Henrik Linusson, Martin Gerdin Wärnberg, Johanna Berg

**Affiliations:** Department of Medical Epidemiology and Biostatistics, Karolinska Institutet, Stockholm, Sweden; Department of Global Public Health, Karolinska Institutet, Stockholm, Sweden; Perioperative Medicine and Intensive Care, Karolinska University Hospital, Solna, Sweden; Center for Research and Innovation, Region Halland, Halmstad, Sweden; School of ITE, Halmstad University, Halmstad, Sweden; Ekkono Solutions AB, Varberg, Sweden; Emergency Medicine, Department of Clinical Sciences, Lund University, Malmö, Sweden

## Abstract

**Background:** Trauma patients present with heterogeneous injury patterns that are challenging to represent in statistical models. Traditional approaches either use high-dimensional one-hot encoding, resulting in sparse features, or aggregate injuries into summary scores that lose patient-specific detail. This study developed data-driven ICD-10 embeddings for trauma injuries and evaluated their ability to preserve injury information.

**Methods:** Using the National Trauma Data Bank, we trained autoencoder models on all trauma patients from 2018 to generate dense vector representations of ICD-10 injury codes. We evaluated embeddings of dimensions 2, 4, 8, 16, and 32 against one-hot encoding using three prediction tasks: in-hospital mortality, emergency department disposition, and blood transfusion within 24 hours. For each hospital included, we trained separate logistic regression and LightGBM models using 2018 data from that hospital, then evaluated performance on 2019 data from the same hospital. Performance was measured using area under the receiver operating characteristic curve (AUC) and stratified by hospital size.

**Results:** In LightGBM models, 8-dimensional embeddings improved AUC compared to one-hot encoding of 0.08 (95% CI: 0.06, 0.10) in small hospitals, 0.03 (0.02, 0.04) in medium hospitals, and 0.02 (0.01, 0.02) in large hospitals, with comparable performance in major hospitals (0.00 [-0.01, 0.01]). In logistic regression, 32-dimensional embeddings showed AUC improvements of 0.03 (0.01, 0.05), 0.02 (0.01, 0.03), and 0.02 (0.02, 0.03) for small, medium, and large hospitals respectively, with similar performance in major hospitals (0.01 [0.00, 0.01]).

**Conclusion:** ICD-10 code injury embeddings with ≥8 dimensions preserve clinically relevant information and can outperform one-hot encoding while reducing dimensionality. The embeddings and software are openly available to support further trauma research and applications.

## 1 Introduction

Trauma comprises physical injury and the body’s response [1]. Trauma patients have diverse injuries, mechanisms, and severities [2]. This heterogeneity complicates representing injuries in a form suitable for machine learning. In clinical practice, injuries are coded with standardised frameworks such as Abbreviated Injury Scale (AIS) and the International Classification of Diseases (ICD) [3, 4]. However, analytical and machine learning pipelines often binarise these codes via one-hot encoding, yielding large, sparse, variable features that discard inherent anatomical context [5, 6].

To keep the feature space manageable, models often aggregate by mechanism, regional severity, or Injury Severity Score (ISS) [7], but such summaries miss patient-specific injury detail, letting clinically distinct patients end up with the same score. Two patients can have the same ISS but very different injuries. Thus, current approaches force a trade-off between high-granularity, high-parameter encodings and low-parameter, low-fidelity summaries, with no scalable method to retain detail without a dramatic increase in features.

Embedding models can represent injuries as dense vectors that capture relationships between injuries. Such representations can support clustering, prediction, and visualisation of trauma patients. However, most prior work in health care has generated embeddings from the structure of coding ontologies, such as ICD-10, rather than from actual patient data. These approaches reproduce how codes are organised and reduce dimensions but do not capture the clinical patterns of how injuries co-occur in trauma patients [5, 8, 9].

This study aimed to develop ICD 10 embeddings of trauma injuries using patient data, evaluate their ability to preserve injury information, and make them available as a resource for future research.

## 2 Materials and Methods

We conducted a retrospective study in two phases: (1) Embedding Model Development and (2) Embedding Model Evaluation. To evaluate whether the embeddings preserved injury information, we tested their performance in downstream prediction tasks relevant to trauma care. We compared embeddings of varying dimensions with one-hot encoding in both linear (logistic regression) and non-linear (LightGBM) models. Predictive performance served as a proxy for information preservation, based on the assumption that accurate predictions require embeddings to retain clinically relevant relationships between injuries. A schematic of the study design is presented in Figure 2.

**Figure 1.**
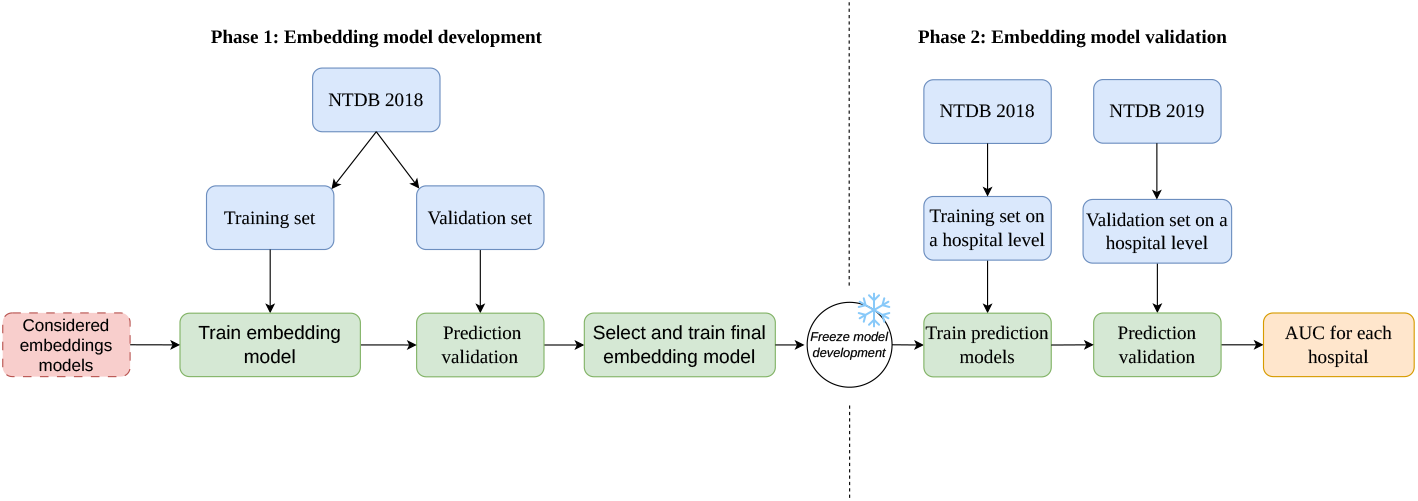
Overview of the two-phase study design: embedding model development and embedding model evaluation.

**Figure 2.**
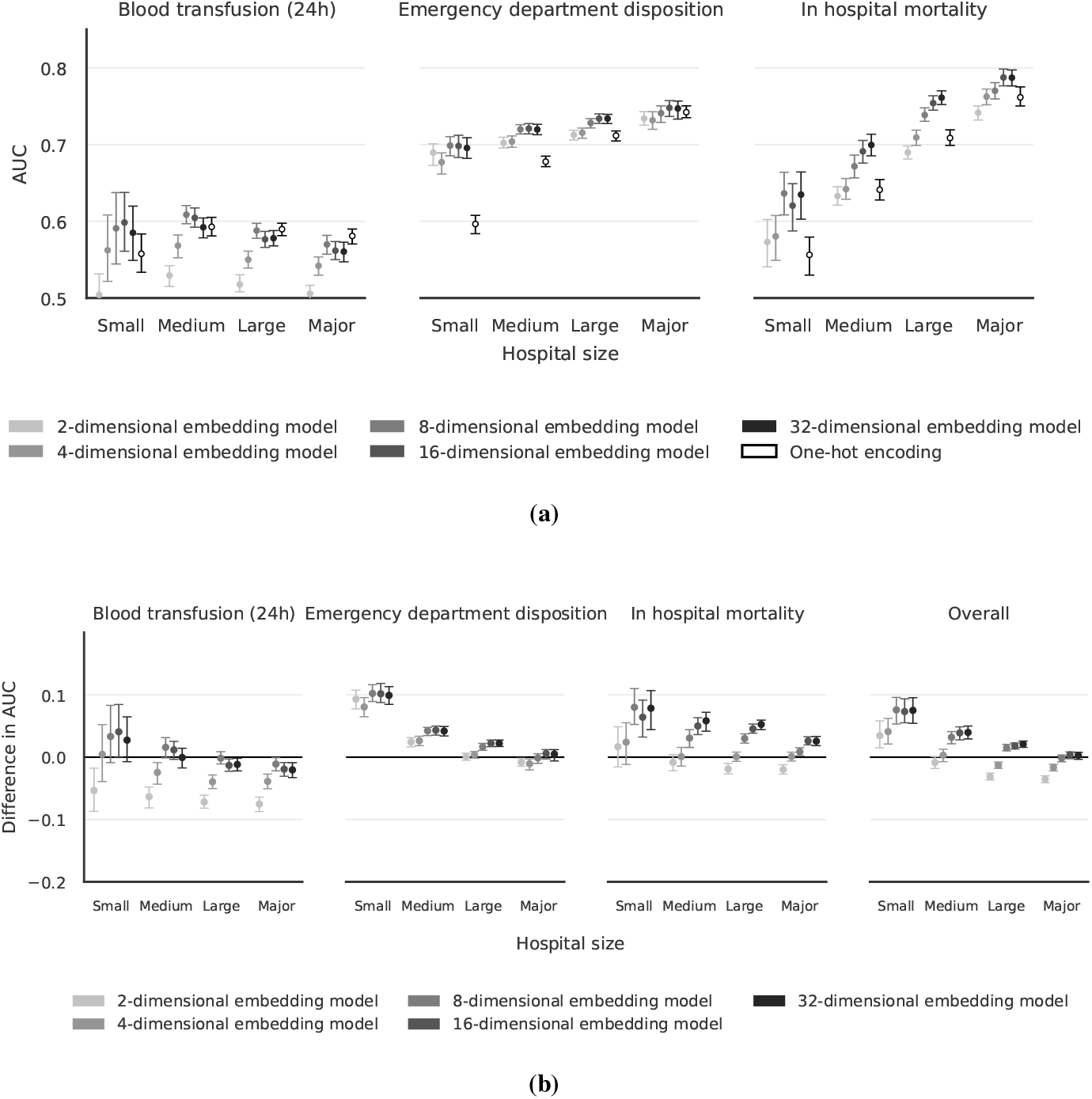
Performance of embeddings and one-hot encoding in LightGBM across hospital sizes. (a) AUC values for different embedding dimensions compared to one-hot encoding for three prediction tasks. (b) AUC differences between embeddings and one-hot encoding, with positive values indicating superior embedding performance.

### 2.1 Data and Participants

In this retrospective registry-based study, we utilised the National Trauma Data Bank (NTDB) maintained by the American College of Surgeons to develop and validate ICD-10 code embeddings for trauma patients [10]. The NTDB was selected for its comprehensive coverage of trauma cases, as it includes patients with trauma ICD-10 diagnosis codes who are admitted, transferred, or die, as well as patients for whom the trauma team is activated regardless of whether ICD-10 codes are present. Additionally, the NTDB provides broad representation of United States trauma hospitals, encompassing Level I–V trauma centres as well as undesignated centres.

To ensure adequate representation of injuries whilst avoiding potential confounding effects from the COVID-19 pandemic, we analysed NTDB data from 1 January 2018 to 31 December 2019. Our study followed a two-phase approach. In the embedding model development phase, data from 2018 were used for training and tuning the embedding models. All patients from this period with non-missing ICD-10 codes were included in the study. For the evaluation phase, data from 2019 served as a held-out evaluation set to evaluate the final embedding model. The evaluation dataset was only used to test the final embedding model, ensuring no data leakage between development and evaluation phases.

### 2.2 Data Preparation

The NTDB uses ICD-10-CM, a clinical modification of the ICD-10, to code conditions with a higher level of granularity. We extracted all ICD-10-CM codes from the NTDB and removed any codes that were not related to injuries. We then translated the ICD-10-CM codes into their corresponding ICD-10 international codes by removing the extraneous granularity that the CM modification adds. This ensures that the model is applicable to datasets that do not use the CM extension. For example, ICD-10-CM code S21.122A (“Laceration with foreign body of the left front wall of the thorax without penetration into the thoracic cavity, initial encounter”) was mapped to the international ICD-10 code S21.1 (“Open wound of the front wall of the thorax”). The code implementing this ICD-10-CM to ICD-10 international translation is available online.

### 2.3 Embedding Model Development

To identify the optimal embedding approach, we evaluated autoencoders; Doc2Vec [11]; dimensionality-reduction of co-occurrence matrices via SVD, UMAP, PCA, and NMF; and BioGPT-based embeddings of ICD-10 descriptions [12]. To preserve the integrity of our 2019 evaluation data, we split the 2018 data into training and test sets for initial model evaluation using a random 70%-30% train-test split.

We trained each candidate embedding model on the 2018 training split and evaluated their performance on the test split using the same downstream prediction tasks planned for the later evaluation. Based on these preliminary evaluations, we selected the vanilla autoencoder architecture for our embedding model architecture as it consistently outperformed the other methods as inputs for the prediction models.

The selected autoencoder architecture was trained using one-hot encoded ICD-10 codes from the NTDB. The model compressed these inputs through a bottleneck layer before reconstructing the original onehot encodings of ICD10 codes. To identify the optimal embedding dimensionality, we trained the model with varying bottleneck dimensions of 2, 4, 8, 16, and 32. We optimised the model using cross-entropy loss and the AdamW optimiser. The hyperparameters that were tuned included network architecture, dropout rates, weight decay, and learning rate. The final model was trained on the complete 2018 dataset. Complete information about the final model can be found in the published source code.

### 2.4 Embedding Model Evaluation

We used one-hot encoding of the ICD-10 codes as a baseline model for comparison with the embeddings. The onehot encoding created a matrix where each row represented a patient and each column represented an ICD-10 code.

To evaluate the embedding model, we compared prediction model performance across several clinically relevant downstream tasks. We compared models using embeddings as features against models using one-hot encoding. For each patient, we generated vector representations using both our embedding model and one-hot encoding of their ICD-10 codes, then used these representations as input features for prediction models. The prediction tasks used three outcomes: in-hospital mortality; emergency department disposition; and blood transfusion within 24 hours. Emergency department disposition was coded as died, home, intensive care unit (ICU), operating room (OR), transferred, ward, or other. The performance on these downstream tasks served only as an internal benchmark against one-hot encoding, rather than benchmarking against other prediction models for these tasks. Both the embeddings and one-hot encoded data were used in a linear classification model (logistic regression) and non-linear classification model (LightGBM), using the default parameters provided by the scikitlearn [13] and LightGBM [14] libraries in Python.

The downstream prediction models were trained at a hospital level, using data from each hospital in NTDB from 2018 and evaluated on data from the same hospital from 2019. Hospitals that did not report during both 2018 and 2019 were excluded. One-hot encoding for each hospital was performed only on codes available in each hospital’s training data; consequently, codes that the one-hot encoding model had not encountered during training were excluded from the evaluation data. The area under the receiver operating characteristic curve (AUC) was used to measure model performance. In multiclass classification tasks, such as disposition from the emergency department, we used the macroaverage AUC (onevsrest). The 95% confidence intervals (CIs) were calculated from nonparametric bootstrapping using 1,000 bootstrap samples.

To evaluate the generalisability of the embeddings across different hospital sizes, we stratified results by hospital size. As the NTDB does not include designated trauma centre levels, we used annual trauma admission volumes as a proxy for hospital size. We analysed the distribution of hospitals in the NTDB (Figure A4), which showed a right-skewed pattern with a mean of 2,894 patients per year. Based on this distribution, we categorised hospitals into four arbitrary groups: small (≤500 patients/year), medium (501–1,250 patients/year), large (1,251–2,500 patients/year), and major (>2,500 patients/year).

## 3 Results

### 3.1 Dataset Characteristics

The study included a total of 2,140,926 trauma patients from the National Trauma Data Bank (NTDB) between 2018 and 2019 (Figure A5). Due to missing ICD-10 codes, 4,972 patients were excluded from the analysis. For the embedding development phase, all patients during 2018 were included, with 1,041,021 patients used to train the embeddings. The embedding evaluation phase required hospitals to be present in both 2018 and 2019 data periods. Due to this requirement, among the 804 hospitals in the NTDB between 1 January 2018 and 31 December 2019, 78 hospitals (10%) were excluded from embedding evaluation (Figure A6). This led to the exclusion of 39,124 patients (2%) from embedding evaluation. The remaining 2,096,830 patients in the embedding model evaluation analysis were split into training (n=1,022,811) and evaluation (n=1,074,019) cohorts for the downstream prediction models based on the year of admission.

The demographic and clinical characteristics were similar between the training and evaluation cohorts for the embedding evaluation phase (Table 1). The majority of patients were male (59%), with a relatively balanced age distribution; the largest group was 25–49-year-olds accounting for 27% of the study population. Most patients presented with mild to moderate injuries, with 50% having an Injury Severity Score (ISS) < 9 and 34% with an ISS between 9–15. The vast majority of patients (92%) had a Glasgow Coma Scale (GCS) score of 13–15. In the distribution of ICD codes per patient, 28% had only a single ICD code documented, while 48% had 3 or more ICD codes. The overall in-hospital mortality rate was 3%. The majority of patients (54%) were admitted directly to a ward from the emergency department, while 18% required ICU admission and 10% needed immediate surgical intervention. Blood transfusion within 24 hours was required for 4% of the trauma patients in the cohort.

**Table 1.** Patient characteristics for the embedding model evaluation analysis split by set.

|  | Train (2018) | Validation (2019) | Overall |
| --- | --- | --- | --- |
| ***n*** | 1,022,811 | 1,074,019 | 2,096,830 |
| **Age, years** |  |  |  |
| 18-24 | 198,620 (21%) | 201,826 (20%) | 400,446 (20%) |
| 25-49 | 266,234 (28%) | 273,773 (27%) | 540,007 (28%) |
| 50-64 | 181,930 (19%) | 189,231 (19%) | 371,161 (19%) |
| 65-79 | 184,879 (19%) | 203,826 (20%) | 388,705 (20%) |
| ≥80 | 125,658 (13%) | 135,115 (13%) | 260,773 (13%) |
| Missing | 65,490 | 70,248 | 135,738 |
| **Sex** |  |  |  |
| Female | 416,211 (41%) | 440,883 (41%) | 857,094 (41%) |
| Male | 606,479 (59%) | 632,983 (59%) | 1,239,462 (59%) |
| Missing | 121 | 153 | 274 |
| **GCS** |  |  |  |
| 3-8 | 54,515 (6%) | 54,361 (5%) | 108,876 (6%) |
| 9-12 | 21,423 (2%) | 22,026 (2%) | 43,449 (2%) |
| 13-15 | 891,101 (92%) | 934,691 (92%) | 1,825,792 (92%) |
| Missing | 55,772 | 62,941 | 118,713 |
| **ISS** |  |  |  |
| <9 (Mild) | 513,193 (50%) | 538,498 (50%) | 1,051,691 (50%) |
| 9-15 (Moderate) | 347,514 (34%) | 369,322 (34%) | 716,836 (34%) |
| 16-25 (Severe) | 95,319 (9%) | 98,056 (9%) | 193,375 (9%) |
| ≥26 (Profound) | 64,188 (6%) | 66,147 (6%) | 130,335 (6%) |
| Missing | 2,597 | 1,996 | 4,593 |
| **Pulse Rate, bpm** |  |  |  |
| <60 | 58,079 (6%) | 62,488 (6%) | 120,567 (6%) |
| 60-100 | 691,203 (69%) | 729,780 (70%) | 1,420,983 (69%) |
| 101-120 | 173,104 (17%) | 179,208 (17%) | 352,312 (17%) |
| >120 | 75,217 (8%) | 76,383 (7%) | 151,600 (7%) |
| Missing | 25,208 | 26,160 | 51,368 |
| **Systolic BP, mmHg** |  |  |  |
| 0 | 6,744 (<1%) | 6,979 (<1%) | 13,723 (<1%) |
| 1-49 | 834 (<1%) | 878 (<1%) | 1,712 (<1%) |
| 50-74 | 6,190 (<1%) | 6,611 (<1%) | 12,801 (<1%) |
| 75-89 | 14,476 (1%) | 15,202 (1%) | 29,678 (1%) |
| ≥90 | 958,086 (97%) | 1,006,559 (97%) | 1,964,645 (97%) |
| Missing | 36,481 | 37,790 | 74,271 |
| **Respiratory Rate, breaths/min** |  |  |  |
| 0 | 5,982 (<1%) | 5,958 (<1%) | 11,940 (<1%) |
| 1-4 | 305 (<1%) | 372 (<1%) | 677 (<1%) |
| 5-9 | 2,758 (<1%) | 2,685 (<1%) | 5,443 (<1%) |
| 10-29 | 943,908 (96%) | 992,954 (96%) | 1,936,862 (96%) |
| ≥30 | 33,012 (3%) | 33,782 (3%) | 66,794 (3%) |
| Missing | 36,846 | 38,268 | 75,114 |
| **Number of ICD codes** |  |  |  |
| 1 | 289,131 (28%) | 304,624 (28%) | 593,755 (28%) |
| 2 | 229,676 (22%) | 242,322 (23%) | 471,998 (23%) |
| 3 | 155,341 (15%) | 162,452 (15%) | 317,793 (15%) |
| 4-5 | 173,185 (17%) | 181,833 (17%) | 355,018 (17%) |
| ≥6 | 175,478 (17%) | 182,788 (17%) | 358,266 (17%) |
| **In hospital mortality** | 35,255 (3%) | 35,724 (3%) | 70,979 (3%) |
| **Emergency department disposition** |  |  |  |
| Died | 9,956 (1%) | 10,156 (<1%) | 20,112 (<1%) |
| Home | 103,755 (10%) | 115,330 (11%) | 219,085 (11%) |
| ICU | 186,589 (19%) | 193,497 (19%) | 380,086 (19%) |
| OR | 107,222 (11%) | 113,540 (11%) | 220,762 (11%) |
| Other | 3,996 (<1%) | 4,436 (<1%) | 8,432 (<1%) |
| Transferred | 36,328 (4%) | 37,279 (4%) | 73,607 (4%) |
| Ward | 546,053 (55%) | 569,056 (55%) | 1,115,109 (55%) |
| Missing | 28,912 | 30,725 | 59,637 |
| **Blood transfusion within 24h** | 36,260 (4%) | 41,728 (4%) | 77,988 (4%) |

Definition of abbreviations: GCS = Glasgow Coma Scale; ISS = Injury Severity Score; BP = Blood pressure; ICD = International Classification of Diseases; ICU = Intensive care unit; OR = Operating room.

The 726 hospitals included in the embedding evaluation analysis were categorised into four size groups based on their yearly patient volume (Table A2): small (n=122, 17%), medium (n=268, 37%), large (n=237, 33%), and major (n=99, 14%). When stratified by hospital size, notable differences emerged in patient characteristics and injury patterns (Table A3). Injury severity varied by hospital size, with major hospitals managing more severe trauma cases (17% with ISS ≥ 16 compared to 9% in small hospitals). The number of ICD codes per patient also increased with hospital size — 40% of patients at small hospitals had only one ICD code, compared to 24% at major hospitals. Conversely, only 8% of patients at small hospitals had six or more ICD codes, compared to 20% at major hospitals. Transfer rates were substantially higher at small hospitals (17%) compared to major hospitals (<1%). In 2018, the mean number of unique ICD-10 codes per hospital was 340 (min: 32, max: 600), corresponding to an average one-hot input dimensionality of 340.

### 3.2 Performance of ICD-10 Embeddings in LightGBM

In the LightGBM model, embeddings with dimensions 8, 16, and 32 demonstrated significantly better performance than one-hot encoding across most hospital sizes for emergency department disposition and in-hospital mortality prediction (Figure 2, Table A4 and A5). The performance advantage was particularly pronounced in smaller hospitals, with improvements gradually diminishing as hospital size increased. For the smaller embeddings (dimensions 2 and 4), results were inconsistent and tended towards inferior performance compared to one-hot encoding, especially in large and major hospitals.

For emergency department disposition specifically, embeddings with dimensions 8, 16, and 32 showed consistent improvements. The 8-dimensional embeddings demonstrated AUC improvements of 0.10 (95% CI: 0.09, 0.12) in small hospitals, 0.04 (95% CI: 0.03, 0.05) in medium hospitals, and 0.02 (95% CI: 0.01, 0.02) in large hospitals compared to one-hot encoding. For major hospitals, embeddings of dimensions 8–32 were similar to one-hot encoding. Similarly, for in-hospital mortality prediction, embeddings sized 8–32 had superior performance. The 8-dimensional embeddings yielded AUC improvements of 0.08 (95% CI: 0.05, 0.11) in small hospitals, 0.03 (95% CI: 0.02, 0.04) in medium hospitals, and 0.03 (95% CI: 0.02, 0.04) in large hospitals. Larger embeddings (16–32 dimensions) showed greater improvements in major hospitals with an AUC difference of 0.03 (95% CI: 0.02, 0.03).

For blood transfusion prediction, results varied by hospital size. The 8-dimensional embeddings demonstrated similar performance to one-hot encoding across all hospital sizes, with AUC differences of 0.03 (95% CI: -0.01, 0.08) in small hospitals, 0.02 (95% CI: 0.00, 0.03) in medium hospitals, 0.00 (95% CI: -0.01, 0.01) in large hospitals, and -0.01 (95% CI: -0.02, 0.00) in major hospitals. Embeddings with dimensions 16 and 32 showed similar results to onehot encoding in small to large hospitals, but inferior results for major hospitals with an AUC difference of -0.02 (95% CI: -0.03, -0.01) for both embedding sizes.

When aggregating results across prediction tasks (Table A6 and A9), the LightGBM model with embeddings of size 8, 16, and 32 demonstrated superior performance in small to large hospitals. Embeddings of dimension 8 had average AUC improvements of 0.08 (95% CI: 0.05, 0.10) for small hospitals, 0.03 (95% CI: 0.02, 0.04) for medium hospitals, and 0.02 (95% CI: 0.01, 0.02) for large hospitals. For major hospitals, these embeddings showed similar results to one-hot encoding with an average AUC difference of 0.00 (95% CI: -0.01, 0.00).

### 3.3 Performance of ICD-10 Embeddings in Logistic Regression

In logistic regression models (Figure 3, Table A7 and A8), embedding performance compared to one-hot encoding varied across prediction tasks. For blood transfusion prediction, embeddings with dimensions 8, 16, and 32 demonstrated substantial improvements across all hospital sizes. The 8-dimensional embeddings yielded the largest improvements, with AUC increases of 0.07 (95% CI: 0.03, 0.11) in small hospitals, 0.08 (95% CI: 0.07, 0.10) in medium hospitals, 0.08 (95% CI: 0.07, 0.09) in large hospitals, and 0.07 (95% CI: 0.06, 0.09) in major hospitals. When utilising smaller dimensional embeddings (dimensions 2 and 4), the models had particularly poor performance across the downstream tasks considered. The gap in performance for these smaller embeddings grew with hospital size.

**Figure 3.**
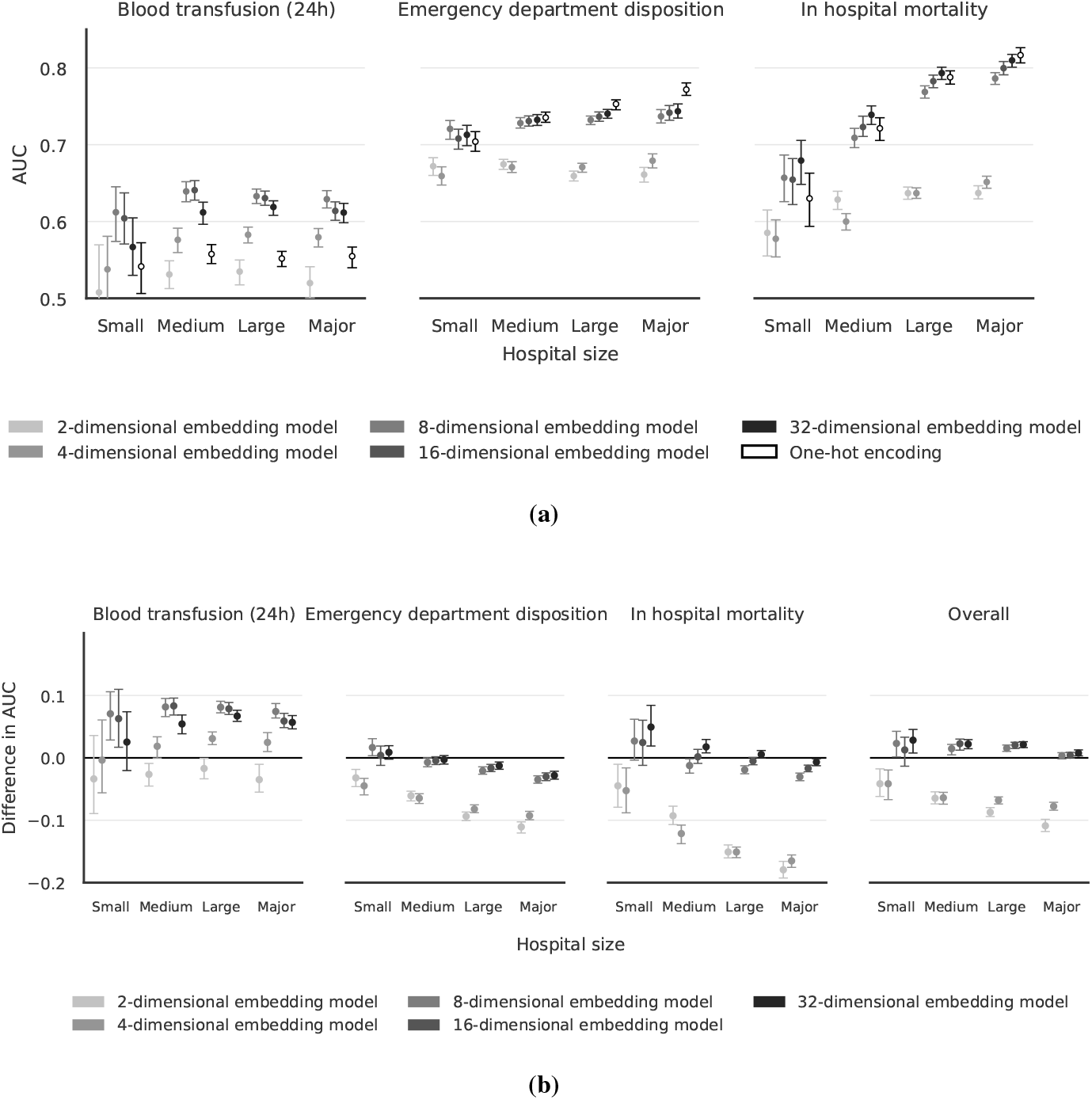
Performance of embeddings and one-hot encoding in logistic regression models across hospital sizes. (a) AUC values for different embedding dimensions compared to one-hot encoding for three prediction tasks. (b) AUC differences between embeddings and one-hot encoding, with positive values indicating superior embedding performance.

For emergency department disposition prediction, embeddings sized 8, 16, and 32 performed similarly to one-hot encoding in small and medium hospitals. However, in large and major hospitals, the embeddings performed worse with AUC differences ranging from -0.01 to -0.03. For in-hospital mortality prediction, 32-dimensional embeddings showed superior performance in small and medium hospitals and similar performance to one-hot encoding in large and major hospitals. The embeddings of dimensions 8 and 16 had similar results to one-hot encoding in small and medium hospitals but inferior results in large and major hospitals, with AUC differences between -0.01 and -0.03.

When aggregating the results for logistic regression models (Table A9), the 32-dimensional embeddings showed superior results for hospital sizes ranging from small to large with improvements of 0.03 (95% CI: 0.01, 0.05), 0.02 (95% CI: 0.01, 0.03), and 0.02 (95% CI: 0.02, 0.03) respectively, and similar results to one-hot encoding for major hospitals with an AUC difference of 0.01 (95% CI: 0.00, 0.01).

## 4 Discussion

We developed ICD-10 embeddings for trauma injuries and found that injury patterns can be represented with substantially fewer dimensions than one-hot encoding. Embeddings with ≥ 8 dimensions preserved clinically relevant information and consistently matched or outperformed one-hot encoding across prediction tasks and modelling approaches, with the largest gains in smaller hospitals. While non-linear models demonstrated acceptable performance with lower dimensions (2 and 4), linear models generally required at least 8 dimensions to be on par with one-hot encoding.

The embedding representation likely encodes clinically meaningful information that is not captured in small samples of one-hot encoded injury codes by modelling latent co-occurrence patterns and relationships between injuries learned from the 1,022,811 patients used to train the embedding model. This advantage is most pronounced in low-volume trauma centres, where direct modelling of individual one-hot codes becomes unreliable due to data sparsity. Furthermore, with one-hot encoding the exact codes present during model training become crucial: even if two codes are in the same ICD-10 chapter, the one-hot encoding model does not inherently recognise their relationship, and codes absent from a hospital’s training data must be excluded when the model is applied to new data. In contrast, embeddings allow prediction models to use the semantic meaning of each code rather than memorising specific codes, enabling generalisation to rare or previously unseen trauma ICD-10 codes by leveraging similarity in the embedding space. The prediction model is therefore not constrained to only use previously encountered trauma ICD-10 codes, but can infer the impact of unseen codes based on their semantic embeddings and similarity to codes it has previously encountered.

Our approach differs from previous methods for representing trauma patients in several key aspects. Compared to traditional methods, e.g. ISS, our method is based on a datadriven approach and can represent injuries in high detail. Furthermore, unlike methods that create embeddings based solely on the hierarchical structure of ICD-10 codes or their textual descriptions [15], our embeddings are derived from real patient data, enabling the discovery of injury relationships observed in clinical practice that are not inherent in the ICD-10 code structure or descriptions. Finally, while previous embedding models for patients exist and display utility, they lack specificity for trauma populations and have more complex input requirements. [5, 8]. Our work addresses this gap by focusing exclusively on representing injuries, because, in trauma, it is crucial to distinguish between the wide variety of injury types and their distinct clinical implications. Furthermore, our model uses a clearly defined input and is restricted to ICD-10 codes.

A key strength of our study is the use of a large, nationally representative trauma database that includes diverse injury patterns across various healthcare centres. This ensured that we could embed the vast majority of trauma ICD-10 codes. The two-phase evaluation approach with temporal separation between development and evaluation cohorts enhances the robustness of our findings. Additionally, our embeddings were designed using base ICD-10 codes rather than the more granular ICD-10-CM codes, improving generalisability across different institutions and healthcare systems.

Some limitations must be acknowledged. The interpretability of embeddings remains challenging compared to traditional one-hot encoding. While embeddings effectively capture relationships between injuries, understanding what the embedding dimensions represent clinically is not straightforward. Additionally, our evaluation was limited to prediction tasks and did not explore other potential applications such as clustering. Finally, while the NTDB represents a diverse population of patients, it is not representative of trauma patients globally. The NTDB represents patients admitted to hospitals in the United States, which may limit the generalisability of our findings to healthcare systems with different injury patterns and coding practices.

There are numerous applications and future research directions for these injury embeddings. We have demonstrated that clinically relevant injury information can be preserved in 8 dimensions. These embeddings could enable trauma patient phenotyping and clustering, potentially revealing clinically meaningful subgroups with distinct prognoses. Additionally, they could be integrated into statistical models to more effectively adjust for trauma phenotypes when evaluating interventions or outcomes. Finally, based on the limitations of the current study, future research should include an external validation using trauma data from healthcare systems outside the United States to ensure generalisability across different injury patterns and coding practices.

## 5 Conclusion

ICD-10 embeddings of trauma injuries with at least 8 dimensions preserve clinically relevant information and, in several prediction tasks, outperform one-hot encoding. By capturing latent relationships between injuries, they provide a more informative representation than isolated codes. The embeddings and accompanying software packages are openly available to enable further research and applications in trauma care.

## Data Availability

All data used in this study are available in the American College of Surgeons National Trauma Data Bank. The code to generate the embeddings, the trained embedding models, and an interface for the models in both Python and R are published online at https://github.com/noacs-io/icd-10-injury-embeddings.

https://github.com/noacs-io/icd-10-injury-embeddings

## Data and Code Availability

All data used in this study are available in the American College of Surgeons National Trauma Data Bank [10]. The code to generate the embeddings, the trained embedding models, and an interface for the models in both Python and R are published online at https://github.com/noacs-io/icd-10-injury-embeddings.

## Ethical considerations

This study used data from the National Trauma Data Bank, which is released in de-identified form under a data use licence following institutional application and fee. Data management was conducted in accordance with the NTDB data use agreement, with no ability to identify individual patients or centres.

## Authors’ Contributions

K.S. and J.B. conceived and designed the study. All authors contributed to the methodology and development of the study protocol. J.B. performed data cleaning and pre-processing of the registry data. K.S. developed the machine learning pipeline and trained the embedding models. K.S. conducted the statistical analyses. K.S. and J.B. analysed and interpreted the study results. K.S. wrote the first draft of the manuscript. M.G.W. secured funding. All authors critically revised the manuscript for important intellectual content and approved the final version. K.S. takes responsibility for the integrity and accuracy of the analysis in this study.

## SUPPLEMENTARY MATERIAL

**Figure A4.**
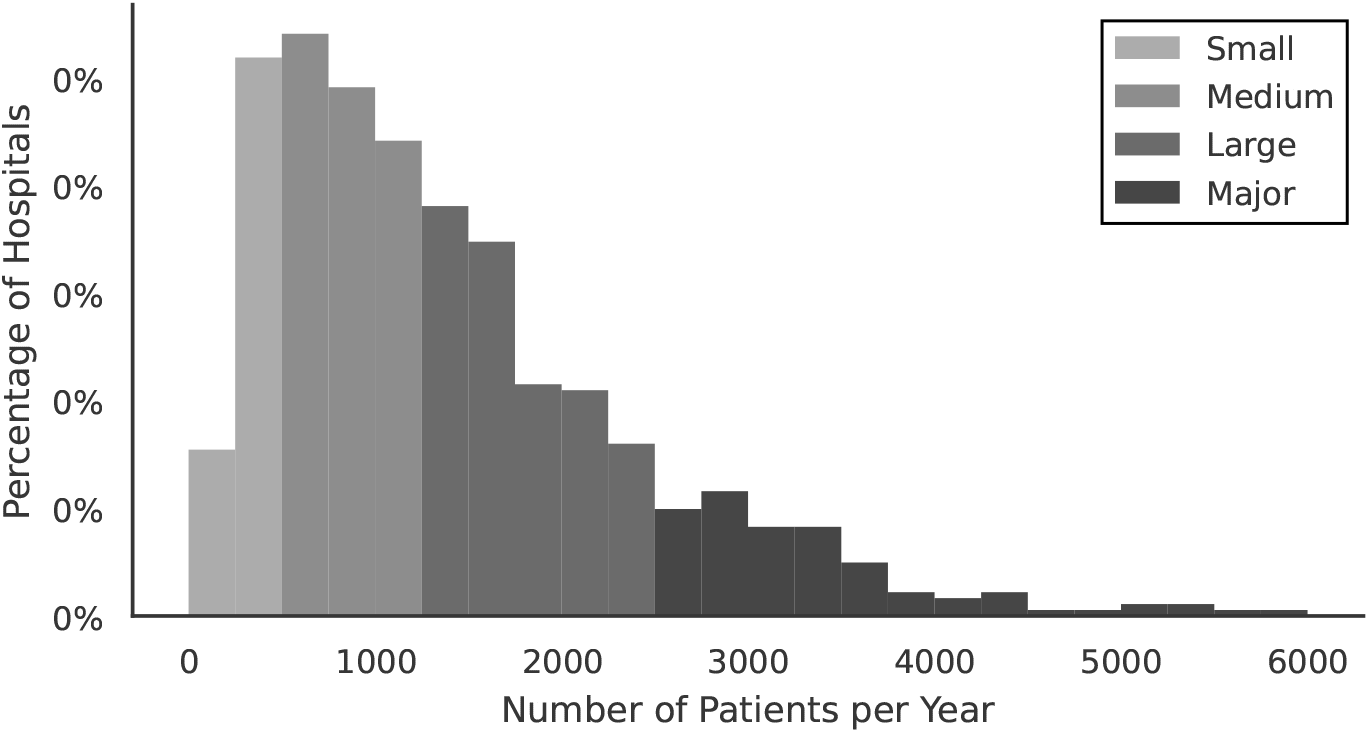
Distribution of trauma hospitals by size category in the NTDB dataset (2018-2019). Hospitals were classified based on annual patient volume.

**Table A2.** Classification of trauma hospitals in the NTDB dataset (2018-2019). Hospitals were categorised into four size groups based on annual patient volume.

| Size Category | Size | n | Percentage |
| --- | --- | --- | --- |
| Small | 0-500 | 122 | 17% |
| Medium | 501-1250 | 268 | 37% |
| Large | 1251-2500 | 237 | 33% |
| Major | >2500 | 99 | 14% |

**Figure A5.**
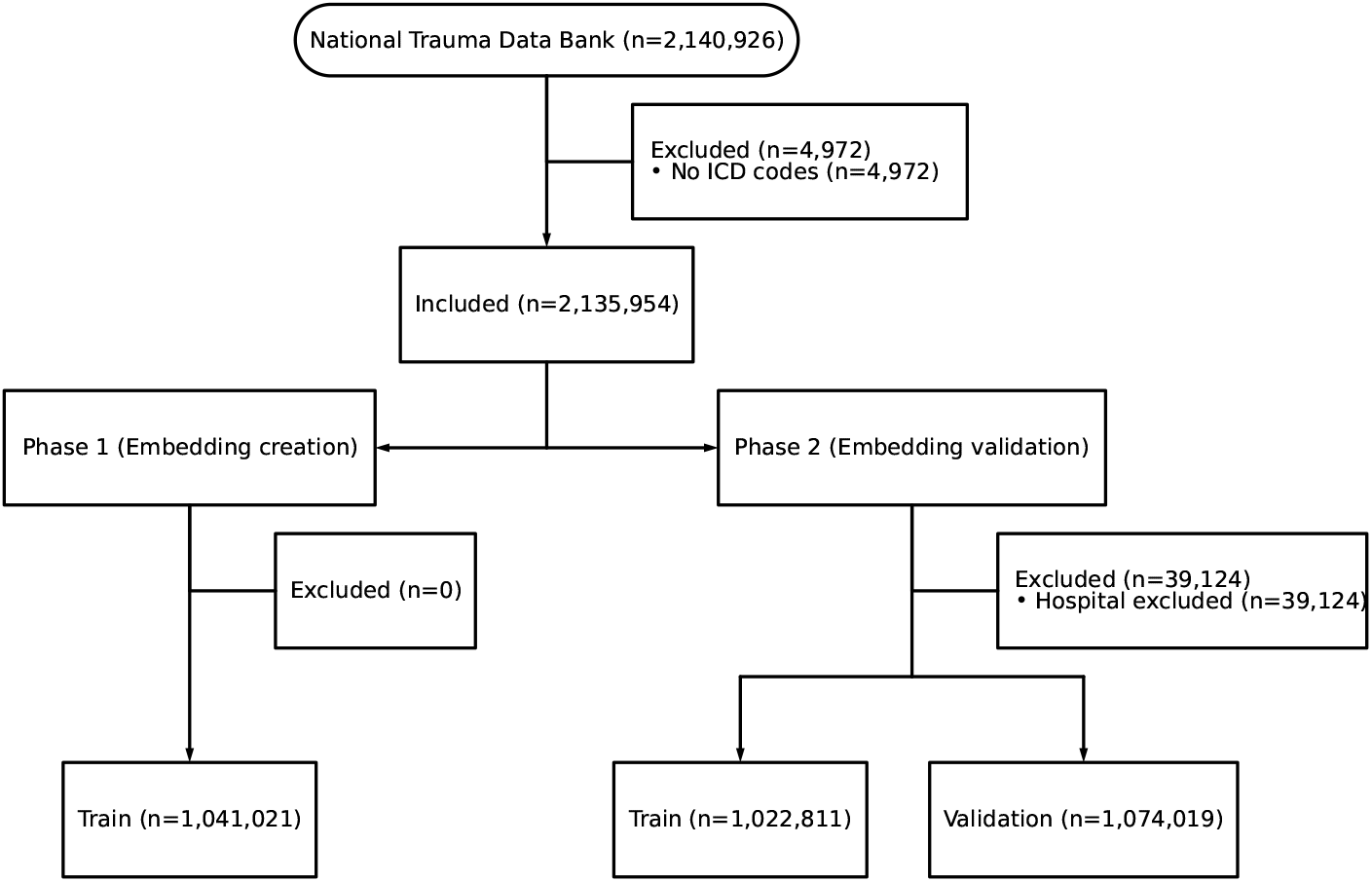
Inclusion flowchart for patients in the study.

**Figure A6.**
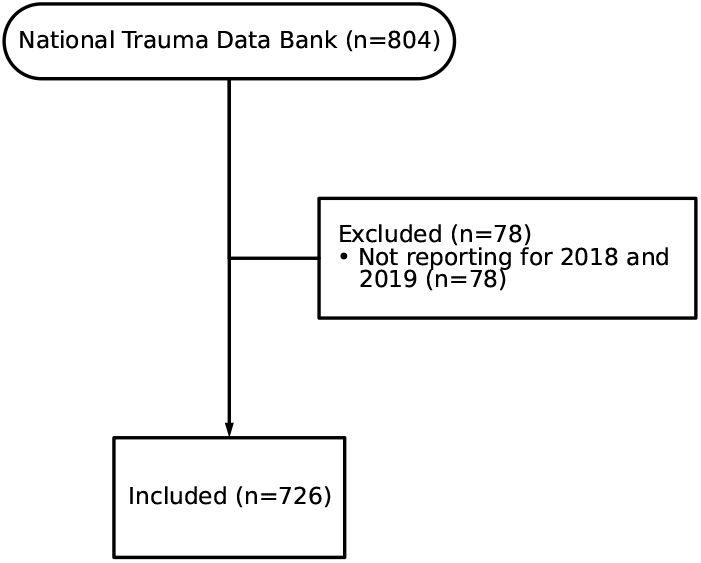
Inclusion flowchart for hospitals in the embedding validation phase.

**Table A3.** Patient characheristics for the embedding validation analysis split by hospital size.

|  | Small | Medium | Large | major |
| --- | --- | --- | --- | --- |
| **n** | 46,403 | 248,634 | 428,774 | 350,208 |
| **Age, years** |  |  |  |  |
| 18-24 | 9,960 (24%) | 49,348 (22%) | 77,840 (19%) | 64,678 (19%) |
| 25-49 | 7,965 (19%) | 50,866 (22%) | 110,867 (28%) | 104,075 (31%) |
| 50-64 | 7,218 (17%) | 40,442 (18%) | 75,850 (19%) | 65,721 (20%) |
| 65-79 | 9,752 (23%) | 50,927 (22%) | 80,796 (20%) | 62,351 (19%) |
| ≥80 | 7,248 (17%) | 37,319 (16%) | 54,569 (14%) | 35,979 (11%) |
| Missing | 4,260 | 19,732 | 28,852 | 17,404 |
| **Sex** |  |  |  |  |
| Female | 21,917 (47%) | 111,586 (45%) | 174,300 (41%) | 133,080 (38%) |
| Male | 24,461 (53%) | 137,027 (55%) | 254,417 (59%) | 217,078 (62%) |
| Missing | 25 | 21 | 57 | 50 |
| **GCS** |  |  |  |  |
| 3-8 | 1,393 (3%) | 9,181 (4%) | 22,262 (5%) | 21,525 (7%) |
| 9-12 | 726 (2%) | 4,345 (2%) | 9,115 (2%) | 7,840 (2%) |
| 13-15 | 39,437 (95%) | 218,051 (94%) | 375,983 (92%) | 301,220 (91%) |
| Missing | 4,847 | 17,057 | 21,414 | 19,623 |
| **ISS** |  |  |  |  |
| <9 (Mild) | 24,762 (54%) | 129,241 (52%) | 212,818 (50%) | 171,677 (49%) |
| 9-15 (Moderate) | 17,016 (37%) | 88,834 (36%) | 148,281 (35%) | 115,191 (33%) |
| 16-25 (Severe) | 2,875 (6%) | 18,597 (7%) | 39,984 (9%) | 36,600 (10%) |
| ≥26 (Profound) | 1,591 (3%) | 11,426 (5%) | 26,884 (6%) | 26,246 (8%) |
| Missing | 159 | 536 | 807 | 494 |
| **Pulse Rate, bpm** |  |  |  |  |
| <60 | 2,859 (6%) | 14,809 (6%) | 25,198 (6%) | 19,622 (6%) |
| 60-100 | 31,113 (69%) | 170,601 (70%) | 292,965 (70%) | 235,101 (69%) |
| 101-120 | 7,407 (16%) | 39,438 (16%) | 71,441 (17%) | 60,922 (18%) |
| >120 | 3,714 (8%) | 17,495 (7%) | 29,731 (7%) | 25,443 (7%) |
| Missing | 1,310 | 6,291 | 9,439 | 9,120 |
| **Systolic BP, mmHg** |  |  |  |  |
| 0 | 192 (<1%) | 1,150 (<1%) | 2,840 (<1%) | 2,797 (<1%) |
| 1-49 | 44 (<1%) | 189 (<1%) | 337 (<1%) | 308 (<1%) |
| 50-74 | 233 (<1%) | 1,356 (<1%) | 2,629 (<1%) | 2,393 (<1%) |
| 75-89 | 613 (1%) | 3,179 (1%) | 5,963 (1%) | 5,447 (2%) |
| ≥90 | 42,958 (98%) | 232,613 (98%) | 403,224 (97%) | 327,764 (97%) |
| Missing | 2,363 | 10,147 | 13,781 | 11,499 |
| **Respiratory Rate, breaths/min** |  |  |  |  |
| 0 | 170 (<1%) | 918 (<1%) | 2,303 (<1%) | 2,567 (<1%) |
| 1-4 | 16 (<1%) | 119 (<1%) | 139 (<1%) | 98 (<1%) |
| 5-9 | 101 (<1%) | 438 (<1%) | 1,123 (<1%) | 1,023 (<1%) |
| 10-29 | 42,565 (95%) | 229,838 (96%) | 397,793 (96%) | 322,758 (96%) |
| ≥30 | 1,741 (4%) | 8,323 (3%) | 13,127 (3%) | 10,591 (3%) |
| Missing | 1,810 | 8,998 | 14,289 | 13,171 |
| **Number of ICD codes** |  |  |  |  |
| 1 | 18,645 (40%) | 84,178 (34%) | 116,599 (27%) | 85,202 (24%) |
| 2 | 11,671 (25%) | 60,046 (24%) | 96,653 (23%) | 73,952 (21%) |
| 3 | 6,333 (14%) | 36,773 (15%) | 65,858 (15%) | 53,488 (15%) |
| 4-5 | 5,746 (12%) | 36,894 (15%) | 74,761 (17%) | 64,432 (18%) |
| ≥6 | 4,008 (9%) | 30,743 (12%) | 74,903 (17%) | 73,134 (21%) |
| **In hospital mortality** | 958 (2%) | 6,583 (3%) | 14,777 (3%) | 13,406 (4%) |
| **Emergency department disposition** |  |  |  |  |
| Died | 360 (<1%) | 1,959 (<1%) | 4,063 (<1%) | 3,774 (1%) |
| Home | 3,964 (9%) | 27,474 (11%) | 40,091 (10%) | 43,801 (13%) |
| ICU | 4,123 (9%) | 35,349 (15%) | 83,649 (20%) | 70,376 (21%) |
| OR | 3,743 (8%) | 21,815 (9%) | 46,798 (11%) | 41,184 (12%) |
| Other | 211 (<1%) | 1,054 (<1%) | 1,876 (<1%) | 1,295 (<1%) |
| Transferred | 7,980 (18%) | 17,282 (7%) | 9,617 (2%) | 2,400 (<1%) |
| Ward | 25,048 (55%) | 134,831 (56%) | 229,453 (55%) | 179,724 (52%) |
| Missing | 974 | 8,870 | 13,227 | 7,654 |
| **Blood transfusion within 24h** | 890 (2%) | 6,936 (3%) | 17,266 (4%) | 16,636 (5%) |

Definition of abbreviations: GCS = Glasgow Coma Scale; ISS = Injury Severity Score; BP = Blood pressure; ICD = International Classification of Diseases; ICU = Intensive care unit; OR = Operating room.

**Table A4.** AUC results for gradient boosted models across different embedding dimensions and hospital sizes for the three selected prediction tasks. The 95% CI is presented within parenthesis.

| Hospital size | One-hot encoding | Embedding (2) | Embedding (4) | Embedding (8) | Embedding (16) | Embedding (32) |
| --- | --- | --- | --- | --- | --- | --- |
| **Blood transfusion (24h)** |  |  |  |  |  |  |
| Small | 0.56 (0.53, 0.58) | 0.5 (0.48, 0.53) | 0.56 (0.52, 0.61) | 0.59 (0.54, 0.64) | **0.6 (0.56, 0.64)** | 0.59 (0.55, 0.62) |
| Medium | 0.59 (0.58, 0.61) | 0.53 (0.52, 0.54) | 0.57 (0.55, 0.58) | **0.61 (0.6, 0.62)** | 0.6 (0.59, 0.62) | 0.59 (0.58, 0.6) |
| Large | **0.59 (0.58, 0.6)** | 0.52 (0.51, 0.53) | 0.55 (0.54, 0.56) | 0.59 (0.58, 0.6) | 0.58 (0.57, 0.59) | 0.58 (0.57, 0.59) |
| Major | **0.58 (0.57, 0.59)** | 0.51 (0.49, 0.52) | 0.54 (0.53, 0.55) | 0.57 (0.56, 0.58) | 0.56 (0.55, 0.57) | 0.56 (0.55, 0.57) |
| **Emergency department disposition** |  |  |  |  |  |  |
| Small | 0.6 (0.58, 0.61) | 0.69 (0.67, 0.7) | 0.68 (0.66, 0.69) | **0.7 (0.69, 0.71)** | 0.7 (0.68, 0.71) | 0.7 (0.68, 0.71) |
| Medium | 0.68 (0.67, 0.68) | 0.7 (0.7, 0.71) | 0.7 (0.7, 0.71) | 0.72 (0.71, 0.73) | **0.72 (0.71, 0.73)** | 0.72 (0.71, 0.73) |
| Large | 0.71 (0.7, 0.72) | 0.71 (0.71, 0.72) | 0.72 (0.71, 0.72) | 0.73 (0.72, 0.73) | 0.73 (0.73, 0.74) | **0.73 (0.73, 0.74)** |
| Major | 0.74 (0.73, 0.75) | 0.73 (0.73, 0.74) | 0.73 (0.72, 0.74) | 0.74 (0.73, 0.75) | **0.75 (0.74, 0.76)** | 0.75 (0.73, 0.76) |
| **In hospital mortality** |  |  |  |  |  |  |
| Small | 0.56 (0.53, 0.58) | 0.57 (0.54, 0.6) | 0.58 (0.55, 0.61) | **0.64 (0.61, 0.66)** | 0.62 (0.59, 0.65) | 0.63 (0.6, 0.66) |
| Medium | 0.64 (0.63, 0.65) | 0.63 (0.62, 0.65) | 0.64 (0.63, 0.66) | 0.67 (0.66, 0.69) | 0.69 (0.68, 0.71) | **0.7 (0.69, 0.71)** |
| Large | 0.71 (0.7, 0.72) | 0.69 (0.68, 0.7) | 0.71 (0.7, 0.72) | 0.74 (0.73, 0.75) | 0.75 (0.74, 0.76) | **0.76 (0.75, 0.77)** |
| Major | 0.76 (0.75, 0.78) | 0.74 (0.73, 0.75) | 0.76 (0.75, 0.77) | 0.77 (0.76, 0.78) | **0.79 (0.78, 0.8)** | 0.79 (0.78, 0.8) |

Definition of abbreviations: AUC = Area Under the Receiver Operating Characteristic Curve; CI = Confidence Interval; ED = Emergency Department.

**Table A5.**
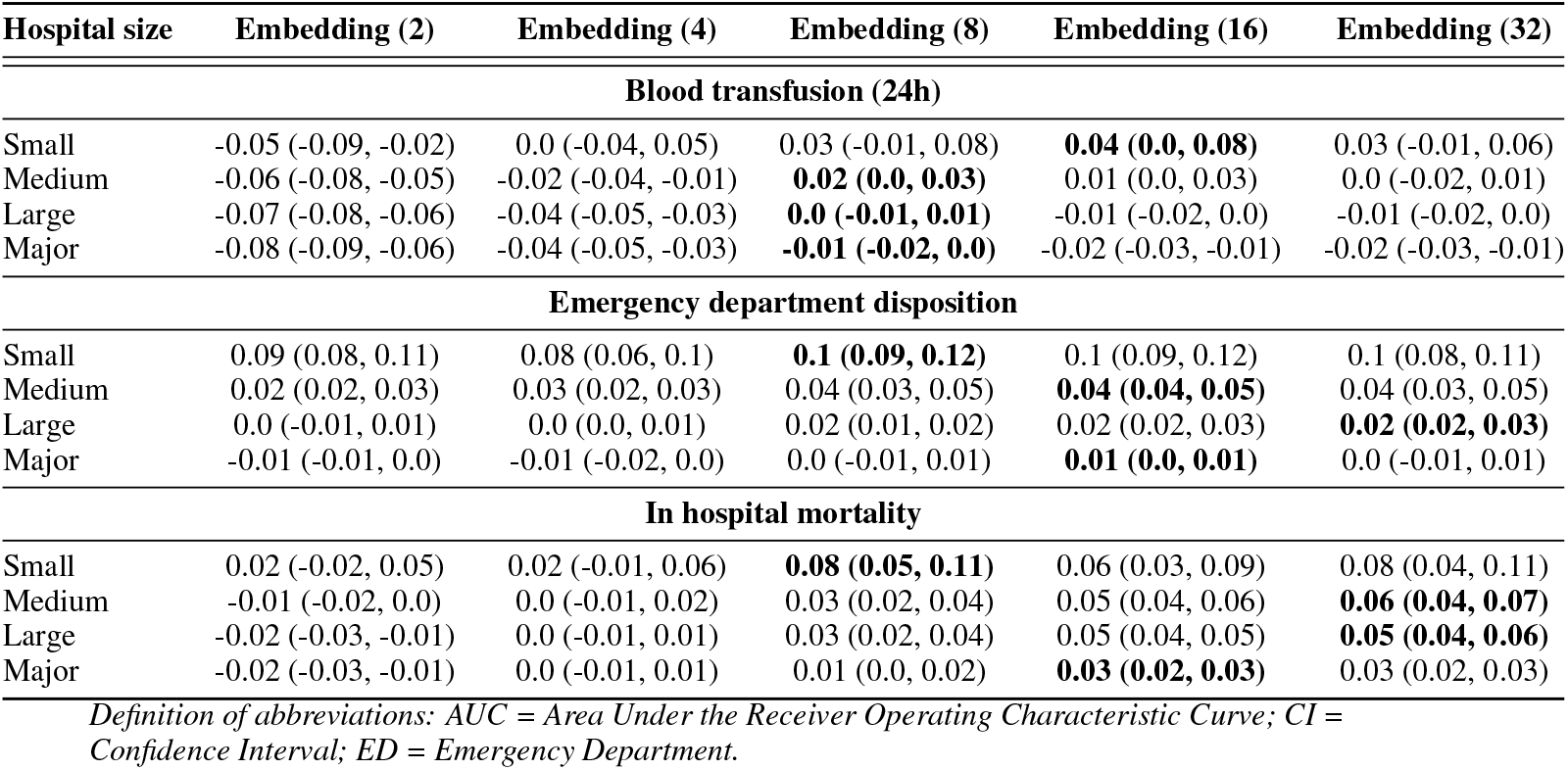
Difference in AUC between embedding models and one-hot encoding for gradient boosted models across the three selected prediction tasks. Positive values indicate superior performance of embeddings compared to one-hot encoding. The 95% CI is presented within parenthesis.

| Hospital size | Embedding (2) | Embedding (4) | Embedding (8) | Embedding (16) | Embedding (32) |
| --- | --- | --- | --- | --- | --- |
| **Blood transfusion (24h)** |  |  |  |  |  |
| Small | -0.05 (-0.09, -0.02) | 0.0 (-0.04, 0.05) | 0.03 (-0.01, 0.08) | **0.04 (0.0, 0.08)** | 0.03 (-0.01, 0.06) |
| Medium | -0.06 (-0.08, -0.05) | -0.02 (-0.04, -0.01) | **0.02 (0.0, 0.03)** | 0.01 (0.0, 0.03) | 0.0 (-0.02, 0.01) |
| Large | -0.07 (-0.08, -0.06) | -0.04 (-0.05, -0.03) | **0.0 (-0.01, 0.01)** | -0.01 (-0.02, 0.0) | -0.01 (-0.02, 0.0) |
| Major | -0.08 (-0.09, -0.06) | -0.04 (-0.05, -0.03) | **-0.01 (-0.02, 0.0)** | -0.02 (-0.03, -0.01) | -0.02 (-0.03, -0.01) |
| **Emergency department disposition** |  |  |  |  |  |
| Small | 0.09 (0.08, 0.11) | 0.08 (0.06, 0.1) | **0.1 (0.09, 0.12)** | 0.1 (0.09, 0.12) | 0.1 (0.08, 0.11) |
| Medium | 0.02 (0.02, 0.03) | 0.03 (0.02, 0.03) | 0.04 (0.03, 0.05) | **0.04 (0.04, 0.05)** | 0.04 (0.03, 0.05) |
| Large | 0.0 (-0.01, 0.01) | 0.0 (0.0, 0.01) | 0.02 (0.01, 0.02) | 0.02 (0.02, 0.03) | **0.02 (0.02, 0.03)** |
| Major | -0.01 (-0.01, 0.0) | -0.01 (-0.02, 0.0) | 0.0 (-0.01, 0.01) | **0.01 (0.0, 0.01)** | 0.0 (-0.01, 0.01) |
| **In hospital mortality** |  |  |  |  |  |
| Small | 0.02 (-0.02, 0.05) | 0.02 (-0.01, 0.06) | **0.08 (0.05, 0.11)** | 0.06 (0.03, 0.09) | 0.08 (0.04, 0.11) |
| Medium | -0.01 (-0.02, 0.0) | 0.0 (-0.01, 0.02) | 0.03 (0.02, 0.04) | 0.05 (0.04, 0.06) | **0.06 (0.04, 0.07)** |
| Large | -0.02 (-0.03, -0.01) | 0.0 (-0.01, 0.01) | 0.03 (0.02, 0.04) | 0.05 (0.04, 0.05) | **0.05 (0.04, 0.06)** |
| Major | -0.02 (-0.03, -0.01) | 0.0 (-0.01, 0.01) | 0.01 (0.0, 0.02) | **0.03 (0.02, 0.03)** | 0.03 (0.02, 0.03) |

Definition of abbreviations: AUC = Area Under the Receiver Operating Characteristic Curve; CI = Confidence Interval; ED = Emergency Department.

**Table A6.** Aggregated mean difference in AUC between embedding models and one-hot encoding for gradient boosted models. Values represent the average AUC difference with 95% CIs across the three prediction tasks, stratified by embedding dimension and hospital size.

| Hospital size | Embedding (2) | Embedding (4) | Embedding (8) | Embedding (16) | Embedding (32) |
| --- | --- | --- | --- | --- | --- |
| Small | 0.03 (0.02, 0.06) | 0.04 (0.02, 0.06) | 0.08 (0.05, 0.1) | 0.07 (0.06, 0.09) | 0.08 (0.05, 0.1) |
| Medium | -0.01 (-0.02, 0.0) | 0.0 (-0.01, 0.01) | 0.03 (0.02, 0.04) | 0.04 (0.03, 0.05) | 0.04 (0.03, 0.05) |
| Large | -0.03 (-0.04, -0.03) | -0.01 (-0.02, -0.01) | 0.01 (0.01, 0.02) | 0.02 (0.01, 0.02) | 0.02 (0.02, 0.03) |
| Major | -0.04 (-0.04, -0.03) | -0.02 (-0.02, -0.01) | 0.0 (-0.01, 0.0) | 0.0 (0.0, 0.01) | 0.0 (0.0, 0.01) |

Definition of abbreviations: AUC = Area Under the Receiver Operating Characteristic Curve; CI = Confidence Interval; ED = Emergency Department.

**Table A7.** AUC results for logistic regression models across different embedding dimensions and hospital sizes for the three selected prediction tasks. The 95% CI is presented within parenthesis.

| Hospital size | One-hot encoding | Embedding (2) | Embedding (4) | Embedding (8) | Embedding (16) | Embedding (32) |
| --- | --- | --- | --- | --- | --- | --- |
| **Blood transfusion (24h)** |  |  |  |  |  |  |
| Small | 0.54 (0.51, 0.57) | 0.51 (0.45, 0.57) | 0.54 (0.49, 0.58) | **0.61 (0.57, 0.65)** | 0.6 (0.57, 0.64) | 0.57 (0.53, 0.6) |
| Medium | 0.56 (0.55, 0.57) | 0.53 (0.51, 0.55) | 0.58 (0.56, 0.59) | 0.64 (0.63, 0.65) | **0.64 (0.63, 0.65)** | 0.61 (0.6, 0.63) |
| Large | 0.55 (0.54, 0.56) | 0.53 (0.52, 0.55) | 0.58 (0.57, 0.59) | **0.63 (0.62, 0.64)** | 0.63 (0.62, 0.64) | 0.62 (0.61, 0.63) |
| Major | 0.55 (0.54, 0.57) | 0.52 (0.5, 0.54) | 0.58 (0.57, 0.59) | **0.63 (0.62, 0.64)** | 0.61 (0.6, 0.63) | 0.61 (0.6, 0.62) |
| **Emergency department disposition** |  |  |  |  |  |  |
| Small | 0.7 (0.69, 0.72) | 0.67 (0.66, 0.68) | 0.66 (0.65, 0.67) | **0.72 (0.71, 0.73)** | 0.71 (0.69, 0.72) | 0.71 (0.7, 0.73) |
| Medium | **0.74 (0.73, 0.74)** | 0.67 (0.67, 0.68) | 0.67 (0.66, 0.68) | 0.73 (0.72, 0.74) | 0.73 (0.72, 0.74) | 0.73 (0.73, 0.74) |
| Large | **0.75 (0.75, 0.76)** | 0.66 (0.65, 0.67) | 0.67 (0.66, 0.68) | 0.73 (0.73, 0.74) | 0.74 (0.73, 0.74) | 0.74 (0.73, 0.75) |
| Major | **0.77 (0.76, 0.78)** | 0.66 (0.65, 0.67) | 0.68 (0.67, 0.69) | 0.74 (0.73, 0.75) | 0.74 (0.73, 0.75) | 0.74 (0.73, 0.75) |
| **In hospital mortality** |  |  |  |  |  |  |
| Small | 0.63 (0.59, 0.66) | 0.59 (0.56, 0.61) | 0.58 (0.55, 0.6) | 0.66 (0.63, 0.69) | 0.65 (0.62, 0.68) | **0.68 (0.65, 0.71)** |
| Medium | 0.72 (0.71, 0.74) | 0.63 (0.62, 0.64) | 0.6 (0.59, 0.61) | 0.71 (0.7, 0.72) | 0.72 (0.71, 0.74) | **0.74 (0.73, 0.75)** |
| Large | 0.79 (0.78, 0.8) | 0.64 (0.63, 0.64) | 0.64 (0.63, 0.64) | 0.77 (0.76, 0.78) | 0.78 (0.77, 0.79) | **0.79 (0.78, 0.8)** |
| Major | **0.82 (0.81, 0.83)** | 0.64 (0.63, 0.65) | 0.65 (0.64, 0.66) | 0.79 (0.78, 0.79) | 0.8 (0.79, 0.81) | 0.81 (0.8, 0.82) |

Definition of abbreviations: AUC = Area Under the Receiver Operating Characteristic Curve; CI = Confidence Interval; ED = Emergency Department.

**Table A8.** Difference in AUC between embedding models and one-hot encoding for logistic regression models across the three selected prediction tasks. Positive values indicate superior performance of embeddings compared to one-hot encoding. The 95% CI is presented within parenthesis.

| Hospital size | Embedding (2) | Embedding (4) | Embedding (8) | Embedding (16) | Embedding (32) |
| --- | --- | --- | --- | --- | --- |
| **Blood transfusion (24h)** |  |  |  |  |  |
| Small | -0.03 (-0.09, 0.04) | 0.0 (-0.06, 0.06) | **0.07 (0.03, 0.11)** | 0.06 (0.02, 0.11) | 0.03 (-0.02, 0.07) |
| Medium | -0.03 (-0.05, -0.01) | 0.02 (0.0, 0.03) | 0.08 (0.07, 0.1) | **0.08 (0.07, 0.1)** | 0.05 (0.04, 0.07) |
| Large | -0.02 (-0.03, 0.0) | 0.03 (0.02, 0.04) | **0.08 (0.07, 0.09)** | 0.08 (0.07, 0.09) | 0.07 (0.06, 0.08) |
| Major | -0.03 (-0.05, -0.01) | 0.02 (0.01, 0.04) | **0.07 (0.06, 0.09)** | 0.06 (0.05, 0.07) | 0.06 (0.05, 0.07) |
| **Emergency department disposition** |  |  |  |  |  |
| Small | -0.03 (-0.05, -0.02) | -0.04 (-0.06, -0.03) | **0.02 (0.0, 0.03)** | 0.0 (-0.01, 0.02) | 0.01 (0.0, 0.02) |
| Medium | -0.06 (-0.07, -0.05) | -0.06 (-0.07, -0.06) | -0.01 (-0.01, 0.0) | 0.0 (-0.01, 0.0) | **0.0 (-0.01, 0.0)** |
| Large | -0.09 (-0.1, -0.09) | -0.08 (-0.09, -0.08) | -0.02 (-0.03, -0.01) | -0.02 (-0.02, -0.01) | **-0.01 (-0.02, -0.01)** |
| Major | -0.11 (-0.12, -0.1) | -0.09 (-0.1, -0.09) | -0.03 (-0.04, -0.03) | -0.03 (-0.04, -0.02) | **-0.03 (-0.03, -0.02)** |
| **In hospital mortality** |  |  |  |  |  |
| Small | -0.04 (-0.08, -0.01) | -0.05 (-0.09, -0.02) | 0.03 (0.0, 0.06) | 0.02 (-0.01, 0.06) | **0.05 (0.02, 0.08)** |
| Medium | -0.09 (-0.11, -0.08) | -0.12 (-0.14, -0.11) | -0.01 (-0.02, 0.0) | 0.0 (-0.01, 0.01) | **0.02 (0.01, 0.03)** |
| Large | -0.15 (-0.16, -0.14) | -0.15 (-0.16, -0.14) | -0.02 (-0.03, -0.01) | -0.01 (-0.01, 0.0) | **0.01 (0.0, 0.01)** |
| Major | -0.18 (-0.19, -0.17) | -0.17 (-0.18, -0.16) | -0.03 (-0.04, -0.02) | -0.02 (-0.02, -0.01) | **-0.01 (-0.01, 0.0)** |

Definition of abbreviations: AUC = Area Under the Receiver Operating Characteristic Curve; CI = Confidence Interval; ED = Emergency Department.

**Table A9.** Aggregated mean difference in AUC between embedding models and one-hot encoding for logistic regression models. Values represent the average AUC difference with 95% CIs across the three prediction tasks, stratified by embedding dimension and hospital size.

| Hospital size | Embedding (2) | Embedding (4) | Embedding (8) | Embedding (16) | Embedding (32) |
| --- | --- | --- | --- | --- | --- |
| Small | -0.04 (-0.06, -0.02) | -0.04 (-0.07, -0.02) | 0.02 (0.0, 0.04) | 0.01 (-0.01, 0.03) | 0.03 (0.01, 0.05) |
| Medium | -0.06 (-0.07, -0.05) | -0.06 (-0.07, -0.06) | 0.01 (0.0, 0.02) | 0.02 (0.01, 0.03) | 0.02 (0.01, 0.03) |
| Large | -0.09 (-0.09, -0.08) | -0.07 (-0.07, -0.06) | 0.02 (0.01, 0.02) | 0.02 (0.01, 0.03) | 0.02 (0.02, 0.03) |
| Major | -0.11 (-0.12, -0.1) | -0.08 (-0.08, -0.07) | 0.0 (0.0, 0.01) | 0.0 (0.0, 0.01) | 0.01 (0.0, 0.01) |

Definition of abbreviations: AUC = Area Under the Receiver Operating Characteristic Curve; CI = Confidence Interval; ED = Emergency Department.

## Notes

### Competing Interest Statement

The authors have declared no competing interest.

### Funding Statement

This study did not receive any funding.

### Author Declarations

This study used data from the National Trauma Data Bank, which is released in de-identified form under a data use licence following institutional application and fee. Data management was conducted in accordance with the NTDB data use agreement, with no ability to identify individual patients or centres. NTDB: https://www.facs.org/quality-programs/trauma/quality/national-trauma-data-bank/

### Summary of Updates

The only change is to update Kelvin Szolnoky's ORCID to the correct one (0000-0002-0554-1872).

## References

[1] Gerdin M. The Risk of Dying: Predicting Trauma Mortality in Urban Indian Hospitals. Karolinska Institutet;.

[2] Michetti CP, Fakhry SM, Brasel K, Martin ND, Teicher EJ, Newcomb A, et al. Trauma ICU Prevalence Project: The Diversity of Surgical Critical Care;4(1):e000288.

[3] Salottolo K, Settell A, Uribe P, Akin S, Slone DS, ONeal E, et al. The Impact of the AIS 2005 Revision on Injury Severity Scores and Clinical Outcome Measures;40(9):999–1003. Available from: https://linkinghub.elsevier.com/retrieve/pii/S0020138309002721.

[4] International Statistical Classification of Diseases and Related Health Problems. 1. 2nd ed.;. .

[5] Steiger E, Kroll LE. Patient Embeddings From Diagnosis Codes for Health Care Prediction Tasks: Pat2Vec Machine Learning Framework;2:e40755.

[6] Stausberg J, Lehmann N, Kaczmarek D, Stein M. Reliability of Diagnoses Coding with ICD-10;77(1):50–7. Available from: https://linkinghub.elsevier.com/retrieve/pii/S1386505606002723.

[7] Zhang T, Nikouline A, Lightfoot D, Nolan B. Machine Learning in the Prediction of Trauma Outcomes: A Systematic Review;80(5):440–55. Available from: https://linkinghub.elsevier.com/retrieve/pii/S0196064422003353.

[8] Miotto R, Li L, Kidd BA, Dudley JT. Deep Patient: An Unsupervised Representation to Predict the Future of Patients from the Electronic Health Records ;6:26094.

[9] Finch A, Crowell A, Bhatia M, Parameshwarappa P, Chang YC, Martinez J, et al. Exploiting Hierarchy in Medical Concept Embedding*;4(1):ooab022. Available from: https://academic.oup.com/jamiaopen/article/doi/10.1093/jamiaopen/ooab022/6172949.

[10] American College of Surgeons. National Trauma Data Bank. American College of Surgeons Committee on Trauma; 2025. Available from: https://www.facs.org/quality-programs/trauma/quality/national-trauma-data-bank/.

[11] Le Q, Mikolov T. Distributed Representations of Sentences and Documents. In: Xing EP, Jebara T, editors. Proceedings of the 31st International Conference on Machine Learning. vol. 32 of Proceedings of Machine Learning Research. Bejing, China: PMLR; 2014. p. 1188–96. Available from: https://proceedings.mlr.press/v32/le14.html.

[12] Luo R, Sun L, Xia Y, Qin T, Zhang S, Poon H, et al. BioGPT: generative pre-trained transformer for biomedical text generation and mining. Briefings in Bioinformatics. 2022 09;23(6). Bbac409. Available from: 10.1093/bib/bbac409.

[13] Pedregosa F, Varoquaux G, Gramfort A, Michel V, Thirion B, Grisel O, et al. Scikit-learn: Machine Learning in Python. Journal of Machine Learning Research. 2011;12:2825–30.

[14] Ke G, Meng Q, Finley T, Wang T, Chen W, Ma W, et al. LightGBM: A Highly Efficient Gradient Boosting Decision Tree. In: Guyon I, Luxburg UV, Bengio S, Wallach H, Fergus R, Vishwanathan S, et al., editors. Advances in Neural Information Processing Systems. vol. 30. Curran Associates, Inc.;. Available from: https://proceedings.neurips.cc/paper_files/paper/2017/file/6449f44a102fde848669bdd9eb6b76fa-Paper.pdf.

[15] Kane MJ, King C, Esserman D, Latham NK, Greene EJ, Ganz DA. A Compressed Language Model Embedding Dataset of ICD 10 CM Descriptions [preprint];. Available from: http://medrxiv.org/lookup/doi/10.1101/2023.04.24.23289046.

